# Application of machine learning and complex network measures to an EEG dataset from DMT experiments

**DOI:** 10.1101/2022.06.14.22276410

**Authors:** Caroline L. Alves, Thaise G. L. de O. Toutain, Joel Augusto Moura Porto, Aruane M. Pineda, Eduardo Pondé de Sena, Francisco A. Rodrigues, Christiane Thielemann, Manuel Ciba

## Abstract

There is a growing interest in the medical use of psychedelic substances as preliminary studies using them for psychiatric disorders have shown positive results. In particularly, one of these substances is N,N-dimethyltryptamine (DMT) an agonist serotonergic psychedelic that can induce profound alterations in state of consciousness.

In this work, we propose a computational method based on machine learning as an exploratory tool to reveal DMT-induced changes in brain activity using EEG data and provide new insights into the mechanisms of action of this psychedelic substance. To answer these questions, we propose a two-class classification based on (A) the connectivity matrix or (B) complex network measures derived from it as input to a support vector machine We found that both approaches were able to automatically detect changes in the brain activity, with case (B) showing the highest AUC (89%), indicating that complex network measurements best capture the brain changes that occur due to DMT use. In a second step, we ranked the features that contributed most to this result. For case (A) we found that differences in the high alpha, low beta, and delta frequency band were most important to distinguish between the state before and after DMT inhalation, which is consistent with results described in the literature. Further, the connection between the temporal (TP8) and central cortex (C3) and between the precentral gyrus (FC5) and the lateral occipital cortex (T8) contributed most to the classification result. The connection between regions TP8 and C3 has been found in the literature associated with finger movements that might have occurred during DMT consumption. However, the connection between cortical regions FC5 and P8 has not been found in the literature and is presumably related to emotional, visual, sensory, perceptual, and mystical experiences of the volunteers during DMT consumption. For case (B) closeness centrality was the most important complex network measure. Moreover, we found larger communities and a longer average path length with the use of DMT and the opposite in its absence indicating that the balance between functional segregation and integration was disrupted. This findings supports the idea that cortical brain activity becomes more entropic under psychedelics.

Overall, a robust computational workflow has been developed here with an interpretability of how DMT (or other psychedelics) modify brain networks and insights into their mechanism of action. Finally, the same methodology applied here may be useful in interpreting EEG time series from patients who consumed other psychedelic drugs and can help obtain a detailed understanding of functional changes in the neural network of the brain as a result of drug administration.

## I. INTRODUCTION

N,N-dimethyltryptamine (DMT) is a substance endogenously produced in various mammals [1], including humans [2], and has serotonin agonist properties. Thus, it is able to bind to serotonin receptors, simulating the neurotransmitter [3]. In [4] it was for the first time suggested that DMT is produced by the pineal gland in stress situations such as birth and death. In [5, 6] it seems clear that it is produced in small quantities by this gland [7]. The substance was first synthesized in 1931 [8], while its psychoactive effects have been described for the first time many years later in 1956 by [9]. When administered externally in large quantities, DMT can cause altered states of consciousness [10], hallucinations [11–13] and spiritual experiences such as communication with ‘presences’ or ‘entities’, plus reflections on death [14]. An exogenous ingestion can be done by smoking or injecting. Its effect by oral ingestion depends on the inhibition of monoamine oxidase, an enzyme that degrades the alkaloid DMT in the liver and intestine [12]. This enzyme and DMT are also found in ayahuasca tea that has been used in the Amazon for a couple of hundred years, being part of the traditional medicine of the natives from this region [15].

Recently, the interest in the medical use of psychedelics has increased significantly. Only last year, in [16] it was identified about 100 psychedelic clinical trials currently being conducted worldwide. This shows an increase in the number of clinical trials, compared to 43 assisted psychedelic therapy clinical trials conducted since 1999. One example is the psychedelic 3,4-methylenedioxymethamphetamine (MDMA) which is already in phase 3 clinical trials for the treatment of post-traumatic stress disorder (PTSD) [17] and major depression with positive results [18]. Another notable example is the psychedelic psilocybin whose therapeutic use in the U.S. has come to be considered a revolutionary therapy for treatment-resistant depression and major depressive disorder [19]. These first promising results suggest that other psychedelic substances, such as lysergic acid diethylamide (LSD), ibogaine hydrochloride, salvia divinorum, 5-MeO-DMT, ayahuasca and DMT, which have been less studied so far, should be investigated in more detail [20].

Only a few studies on administration of micro- or low-dose DMT to non-human species (predominantly rats) have been published in the scientific literature [21]. In [22], a low dose of DMT was administered to rats resulting in changes in frequency and amplitude of spontaneous excitatory postsynaptic currents (EPSCs) in the prefrontal cortex (PFC) that lasted long even after the drug was removed from the body. In [23], it was described that chronic, intermittent, low doses of DMT produced an antidepressant effect and increased fear extinction learning in rats without affecting working memory or social interaction. For a high dose of DMT (10 mg/kg), an increase in the density of the dendritic spines in the prefrontal cortex was found in rodents and antidepressant and anxiolytic behavioral effects were observed [24]. In humans, a single dose of 0.1 mg/kg of DMT caused an apparent anxiolytic effect shown in [25]. Other studies using inhaled 5-MeO-DMT also observed complete mystical experiences in 75% of volunteers [26] and improvements in depression and anxiety, which were associated with greater intensity of mystical experiences, with spiritual and personal meaning of the experience, when using this substance [27].

Summarizing, there is some evidence that DMT may be effective for the treatment of depression and post-traumatic stress disorder. However, most studies have been conducted in animals and thus only have a reduced power. Therefore, more in-depth studies on DMT, its mechanisms in the brain, and its potential clinical effects in humans are needed, since there are few studies investigating the use of DMT in humans through EEG [28–30] and fMRI [31].

The application of mathematical methods of graph theory yielded interesting insights into the complex network structure of the human brain. It is known from the literature [32–37] that the topology of brain is a small world network. Networks of this type combine completely random structural characteristics and regular connection topologies. They also preserve a high degree of connectivity between local neighborhoods, while allowing all their nodes ^1^ which to be connected to surprisingly short paths [32]. Altering this topology is also associated with pathological states [38–41] and the use of substances such as psychedelics [42–44]. Notably, complex networks parameters have been used as biomarker for several diseases [45, 46]. The use of complex networks is widely used in EEG to characterize the functional networks of the Brain [47–50].

In this context, machine learning (ML) has been used for more accurate and automatic medical diagnosis [51–58]. Compared to traditional statistical techniques, this approach has the advantage of not relying on prior assumptions (such as adequate distribution, independence in observations, absence of multicollinearity, and interaction problems) and, moreover, are suited to automatically analyze and capture non-linear complex relationships in data[59, 60]. As brain data are characterized by high complexity and highly correlated brain regions, ML algorithms have been widely used as a important tool capable of detecting acute and permanent abnormalities in the brain [61–63]. On the other hand, ML shows a lack of interpretability and a black box nature that is an especially disadvantageous general limitation when it comes to understanding medical data [64, 65]. In the last years new techniques have emerged to help in the interpretation of machine learning results. Most notable is the SHapley Additive ExPlanations (SHAP)values method [66]. This metric enables the identification and prioritization of features and can be used with any machine learning algorithm [67–69].

The present work aims to investigate EEG data using ML as an exploratory tool to detect temporal changes in the brain functionality of participants after DMT consumption. The study raised the following research questions:

- Can we automatically detect changes in the functional network structure induced by DMT using ML ?
- Which new insights into the mechanisms of action of DMT can we draw when we use ML in combination with SHAP values?

To answer these questions, we propose a two-class classification based on (A) the connectivity matrix or (B) complex network measures derived from it as input to a support vector machine (SVM) [70]. SVM has been used with excellent results for the classification of complex network measures before [50, 71, 72]. Furthermore, this ML algorithm can handle problems where the sample size of the data is generally smaller in comparison to the dimensionality of its feature space and is therefore applicable to the study of brain disorders with neuroimaging [73], whose data have these characteristics, and also this is the case of the data in this work (the case A with connectivity matrix).

For a biological interpretation the DMT-induced changes, the SHAP values method was applied to identify the features that contributed most to the classification (feature ranking). A robust workflow has been developed usable for medical professionals who are interested in the interpretation of brain network modifications due to DMT (or other psychedelics).

## II. DATA

The data used for this study has been published in [74] and is public available in a raw format ^2^. Thirty-five healthy male and female subjects (7 women and 28 men), volunteered to inhale, using pipes, 40 mg of free DMT extracted from the root of Mimosa hostilis. It should be noted that all participants had previous experiences with ayahuasca. Recordings were made with 24 electrodes, following the EEG electrode positions in the standard 10 – 20 location system. These channels are: Fp1, Fp2, Fz, F7, F8, FC1, FC2, Cz, C3, C4, T7, T8, CPz, CP1, CP2, CP5, CP6, TP9, TP10, Pz, P3, P4, O1, and O2. The recordings on the subjects started 10 minutes before DMT inhalation, 5 min with eyes closed and 5 minutes with eyes open. After DMT use, subjects were recorded about 6 min (6 ± 1.4 min).

## III. METHODOLOGY

In an earlier work of the authors [75], ML in combination with complex network measures was successfully applied to EEG data recorded after ayahuasca consumption to detect changes in brain activity. For this purpose, different levels of data abstraction were used as input: (a) the raw EEG time series, (b) the correlation of the EEG time series, and (c) the complex network measures calculated from (b). Several ML algorithms were tested and the best performance was obtained with the SVM at the abstraction levels (b) and (c). Based on this result, we decided to use in the present work connectivity matrices (see subsection III B) and derived complex network measures (see subsection III C as input for a SVM).

More details are displayed in Figure 1 which summarizes the methodology workflow. In short, EEG time series were separated by filtering in eight frequency bands. In a next step, preprocessing of the EEG time series were performed to obtain the connectivity matrices for each frequency band (and the unfiltered signal), see Figure 1-A and B with details of this process described in subsection III A and III B. In a second step, complex networks measures are derived from the connectivity matrices as described in subsection III C, see Figure 1-C and both types of data sets were used as input to a SVM as described in subsection III D. For interpretation of the classification results the feature ranking algorithm SHapley Additive exPlanations (SHAP) is finally applied as described in subsection III E.

**FIG. 1:**
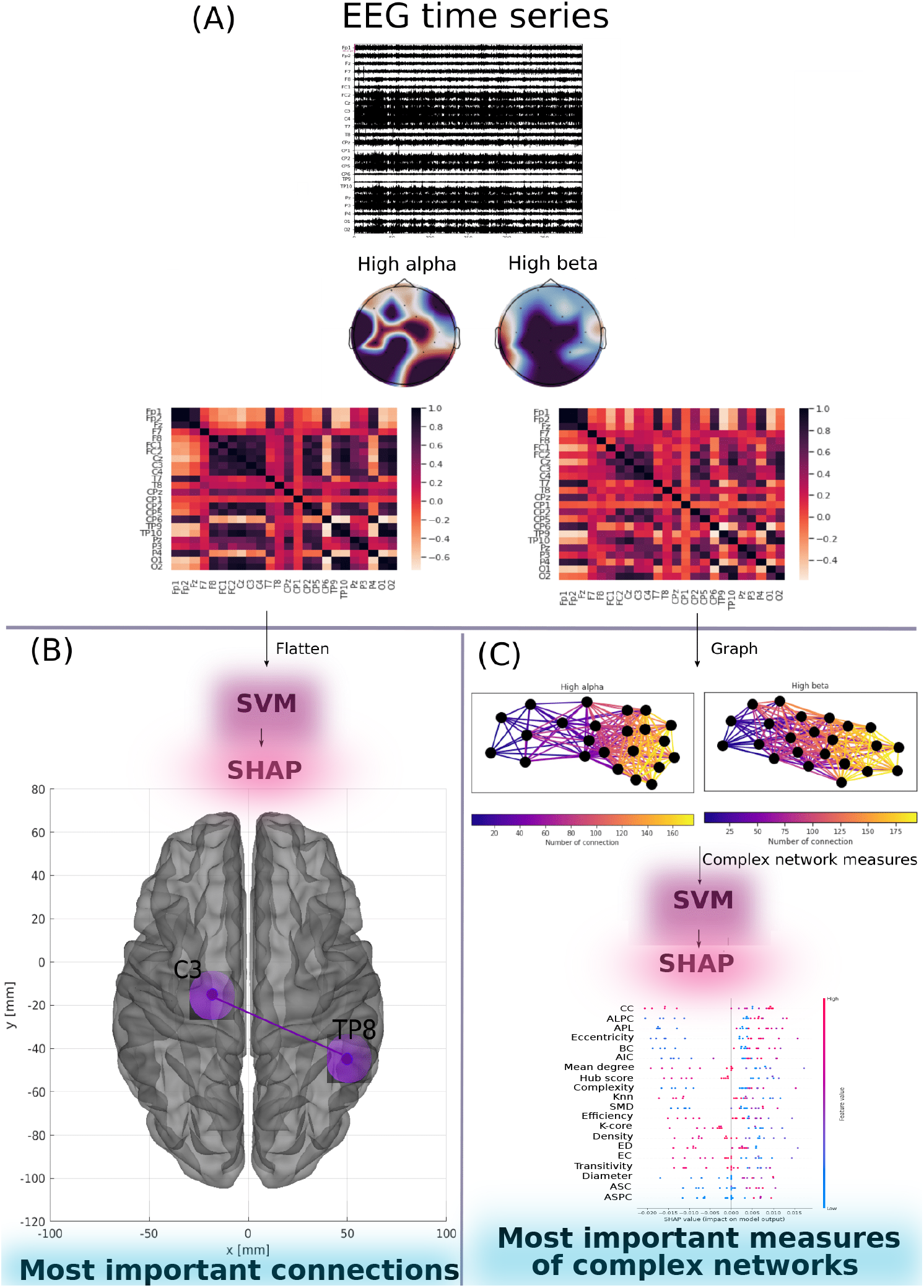
Diagram showing the methodology used in the present work. In **(A) Data preprocessing**, described in the subsection III A, the EEG time series are filtered to remove artifacts (in the picture the time series of a subject at the time the DMT was used) and then separated into the frequency bands high alpha, low alpha, high beta, mid-range beta, low beta, gamma, delta and theta (as an example in the picture the topographic map for the frequencies high alpha and high beta for the same subject). For each band the correlation between the channels is calculated using Pearson’s correlation to obtain a 24×24 connectivity matrix. In **(B) Connectivity matrices**, described in the subsection III B, where the connectivity matrices are flattened into a vector that are put into the SVM in order to verify the most important connections with the use of DMT (in the figure the best performing model, using the high alpha, low beta and delta bands, found TP8 and C3 as the main connections). In **(C) complex network measures**, described in the subsection III C, where the connectivity matrices are analyzed as graphs (in the figure for the same subject, the graph for the frequencies high alpha and high beta, where the number of connections in each node varies according to the color bar) and from them are extracted measures of complex networks that are applied in the SVM and the best model found for the delta frequency found the closeness centrality as the main measure.

### A. Data preprocessing

First, a high-pass filter with a cut off frequency of 0.5 Hz was used to remove artifacts such as electrogalvanic signals and motion artifacts [76]. This type of filtering is widely used in the literature [77–80]. To remove eye artifacts, we employed an independent component analysis (ICA) approach in which EEG signals are decomposed to maximize independent components and those with eye activity are identified and eliminated [81]. An example of the ICA analysis for a subject using DMT can be seen in Figure 2. This analysis was done with a python package called MNE [82] using an algorithm based on maximum information (Infomax) perspective [83].

**FIG. 2:**
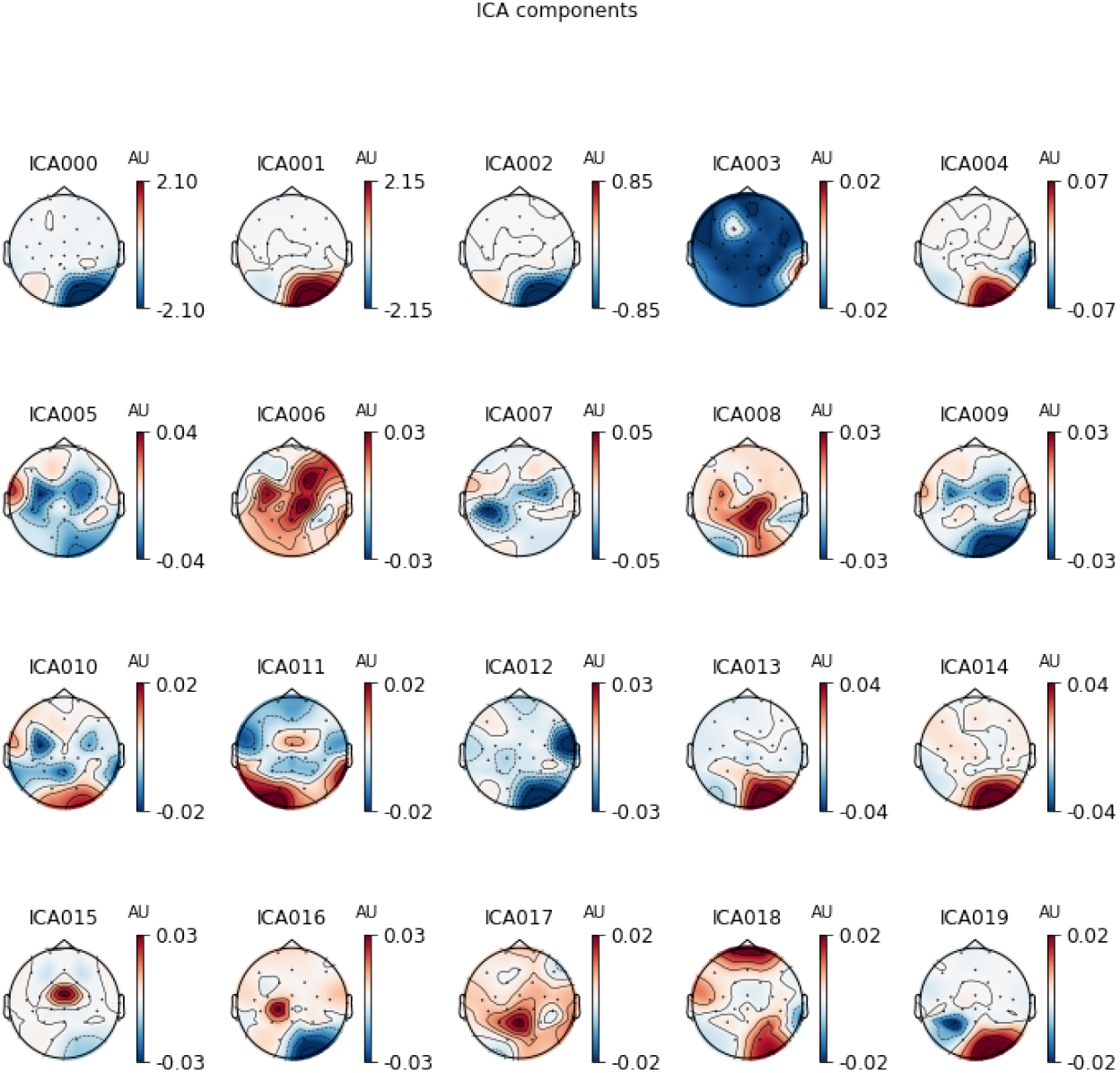
Scalp topography maps generated by ICA analytics recorded from a subject shortly after inhalation of DMT. The EEG signal was decomposed into twenty principal components. The component with ocular activity (see ICA018 with activity in the frontal region near the eyes) is removed and an artifact-free EEG signal is reconstructed.

In the next step, EEG time series were separated by filtering in eight frequency bands: high alpha (10-12 Hz), low alpha (8-10 Hz), low beta waves (12–15 Hz), mid-range beta waves (15–20 Hz), high beta waves (18–40 Hz), gamma (30 - 44 Hz), delta (0.5 – 3 Hz) and theta (4 – 7 Hz).

### B. Connectivity matrices

Connectivity matrices were calculated by the well-known Pearson correlation. It is a widely used and successfully approved measure to capture the correlation of EEG electrodes [84–89]. The Pearson correlation was calculated for all electrode pairs and all frequency bands (including the unfiltered signal). The connectivity matrices serve as input for the following steps as described in subsections III B and III C. To input the data into the ML algorithm, the connectivity matrices were flattened into one vector. Then, all vectors were sequentially merged into a 2D matrix where each column represents a connection between two electrodes and each row represents a subject. Such 2D matrices were generated for each frequency band (and the unfiltered signal).

### C. Complex network measures

For each connectivity matrix, a complex network graph was generated to extract different measures. To input the data into the ML algorithm, the complex network measures were stored in a matrix, where each column represents a complex network measure and each row a subject. Such a 2D matrices were generated for all frequencies bands (and the unfiltered signal).

The following complex network measures were calculated: Assortativity [90, 91], average path length (APL) [92], betweenness centrality (BC) [93], closeness centrality (CC) [94], eigenvector centrality (EC) [95], diameter [96], hub score [97], average degree of nearest neighbors [98] (Knn), mean degree [99], second moment degree (SMD) [100], entropy degree (ED) [101], transitivity [102, 103], complexity, k-core [104, 105], eccentricity [106], density [107], and efficiency [108]. In addition, newly developed metrics (described in detail in [75]) reflecting the number of communities in a complex network are applied. Community detection (also called clustering graph) is one of the fundamental analyses of complex networks aiming to decompose the network in order to find densely connected structures, so-called communities [109–111]. However, the community detection measures need to be transformed into a single scalar value to include them in the matrix. To this aim, we perform the community detection algorithms to find the largest community, then calculate the average path length within this community and receive a single value as the result. The community detection algorithms used were: Fastgreedy community (FC) [112], infomap community (IC) [113], leading eigenvector community (LC) [114], label propagation community (LPC) [115], edge betweenness community (EBC) [116], spinglass (SPC) [117], and multilevel community (MC) [118]. To indicate our approach, we extended the given abbreviations by the letter “A” (for average path length) as follows: AFC, AIC, ALC, ALPC, AEBC, ASPC, and AMC

### D. Machine learning process

In order to classify these two levels of data abstraction, namely the connectivity matrix and the complex network measures, the matrices were sampled by separating them into training (train) and test sets, with 25% of the data composing the test set. Then, for a reliable model, a k-cross validation was used [119], with k = 10 (value widely used in the literature [120–124]). For the training process, the training sets were applied to the SVM. SVM is based on the search for a hyperplane that geometrically divides samples into two classes. Three important hyper-parameters of the SVM have been considered in this work:

- Kernel function: also known as kernel trick, has the function of projecting the input vectors in higher dimensions, because by increasing the dimension of the problem, the probability of it becoming a linearly separable problem increases, which makes it easier to solve [125, 126].
- Regularization parameter C: this is the penalty term of the optimization problem and is an added constant that creates flexible margins with respect to the optimal hyperplane found.
- Gamma: defines how much influence a single training example has. When the gamma value is too small, the model is too restricted and fails to capture the complexity of the data.

To find the best parameters, these hyper-parameters were optimized with the grid search method, widely used in the literature [127–131]. The grid search combines in a comprehensive way all values of the parameters selected for the models using some metrics to evaluate the performance of these combinations, which in the present work was the area under ROC curve (AUC) (for explanation see below). Here, we used the following functions as values for the kernel: gaussian (rbf), polynomial (poly), sigmoid and linear. Optimized values for parameter C and gamma are displayed in Appendix A.

For evaluation, the standard performance metrics accuracy (Acc.) was used as described in [132–136]. As we have a two-class (negative and positive) classification problem, other metrics like Precision and Recall are considered, also common in the literature [137–140]. Precision (also called specificity) corresponds to the hit rate in the negative class (here corresponding no effect induced by DMT). Whereas Recall (also called sensitivity) measures how well a classifier can predict positive examples (hit rate in the positive class), here related with an effect of DMT. Another well-known measure, see [128, 141, 142], is the F1 score which is the harmonic mean of the recall and precision [143]. For visualization of these two latter measures, the receiver operating characteristic (ROC) curve is a common method as it displays the relation between the rate of true positives and false positives. The area below this curve, called area under ROC curve (AUC) has been widely used in classification problems [130, 132, 144, 145]. The value of the AUC varies from 0 to 1, where the value of one corresponds to a classification result free of errors. *AUC* = 0.5 indicates that the classifier is not able to distinguish the two classes equal to the random choice. Furthermore, we consider the micro average of ROC curve, which computes the AUC metric independently for each class (calculate AUC metric for healthy individuals, class zero, and separately calculate for unhealthy subjects, class one) and then the average is computed considering these classes equally. The macro average is also used in our evaluation, which does not consider both classes equally, but aggregates the contributions of the classes separately and then calculates the average.

### E. Feature Ranking

As described in I, most notable technique for interpreting ML results is the SHAP values method based on the Shapley value concept which has its origin in game theory [146, 147], where it aims to assign payoffs to players depending on their contribution to the total payoff in the game. In addition, those who cooperate in a coalition receive a certain profit from this cooperation [148]. Applying this approach to our ML problem, each feature corresponds to a player in a game and the prediction corresponds to the payoff. Thus Shapley’s values tell us how to distribute the payoff fairly among the features [149].

Here, we used this methodology to evaluate which complex network measures and which correlation between electrodes (brain regions) contributed most to the classification result allowing for a biological interpretation of the results obtained with our ML algorithms.

## IV. RESULTS

ML was applied for two different levels of data abstraction: (A) the correlation of EEG time series (connectivity matrix) and (B) the complex network measures calculated from (A). We found that both approaches were able to automatically detect acute changes in the brain activity induced by the inhalation of DMT. The highest classification performance was obtained for the complex network measures with an AUC of 89% (see Table I). The following subsections IV A and IV B describe the results in more detail.

**TABLE I:**
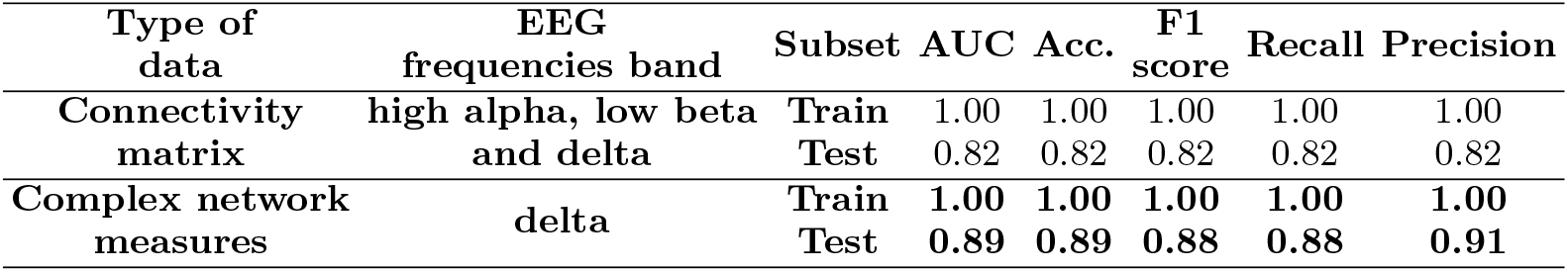
Performances of the SVM classifier. The classification of complex network measures capture the changes in the brain due to DMT slightly better than the connectivity matrix. Best performance is highlighted in bold.

| Type of data | EEG frequencies band | Subset | AUC | Acc. | F1 score | Recall | Precision |
| --- | --- | --- | --- | --- | --- | --- | --- |
| Connectivity matrix | high alpha, low beta and delta | Train | 1.00 | 1.00 | 1.00 | 1.00 | 1.00 |
|  |  | Test | 0.82 | 0.82 | 0.82 | 0.82 | 0.82 |
| Complex network measures | delta | Train | **1.00** | **1.00** | **1.00** | **1.00** | **1.00** |
|  |  | Test | **0.89** | **0.89** | **0.88** | **0.88** | **0.91** |

### A. Connectivity matrix

EEG data recorded from subjects before DMT inhalation (control with eyes closed) and those after inhalation of DMT were filtered and divided into eight frequency bands as described in III A. Detailed results for each frequency band are given in the appendix B. The best performance was achieved for the low beta frequency band (test sample performance with mean AUC of 0.78, mean precision of 0.78, mean F1 score of 0.78, mean recall of 0.78, and mean Acc. of 0.78) followed by the high alpha and delta frequency bands (test sample performance for the both frequency band was a mean AUC of 0.72, mean precision of 0.72, mean F1 score of 0.72, mean recall of 0.72, and mean Acc. of 0.72). Clearly better results were achieved by combining these frequency bands whose test sample performance a mean AUC of 0.82, mean precision of 0.82, mean F1 score of 0.82, mean recall of 0.82, and mean Acc. of 0.82. Furthermore, see appendix D for similarity of results obtained for each frequency.

In Figure 3, the confusion matrix (3-(a)), the learning curve (Figure 3-(b)), and the ROC curve (3-(c)) are displayed. The learning curve evaluates the predictability of the model by varying the size of the training set [69]. Results show that the entire database is required to achieve the highest validation accuracy.

**FIG. 3:**
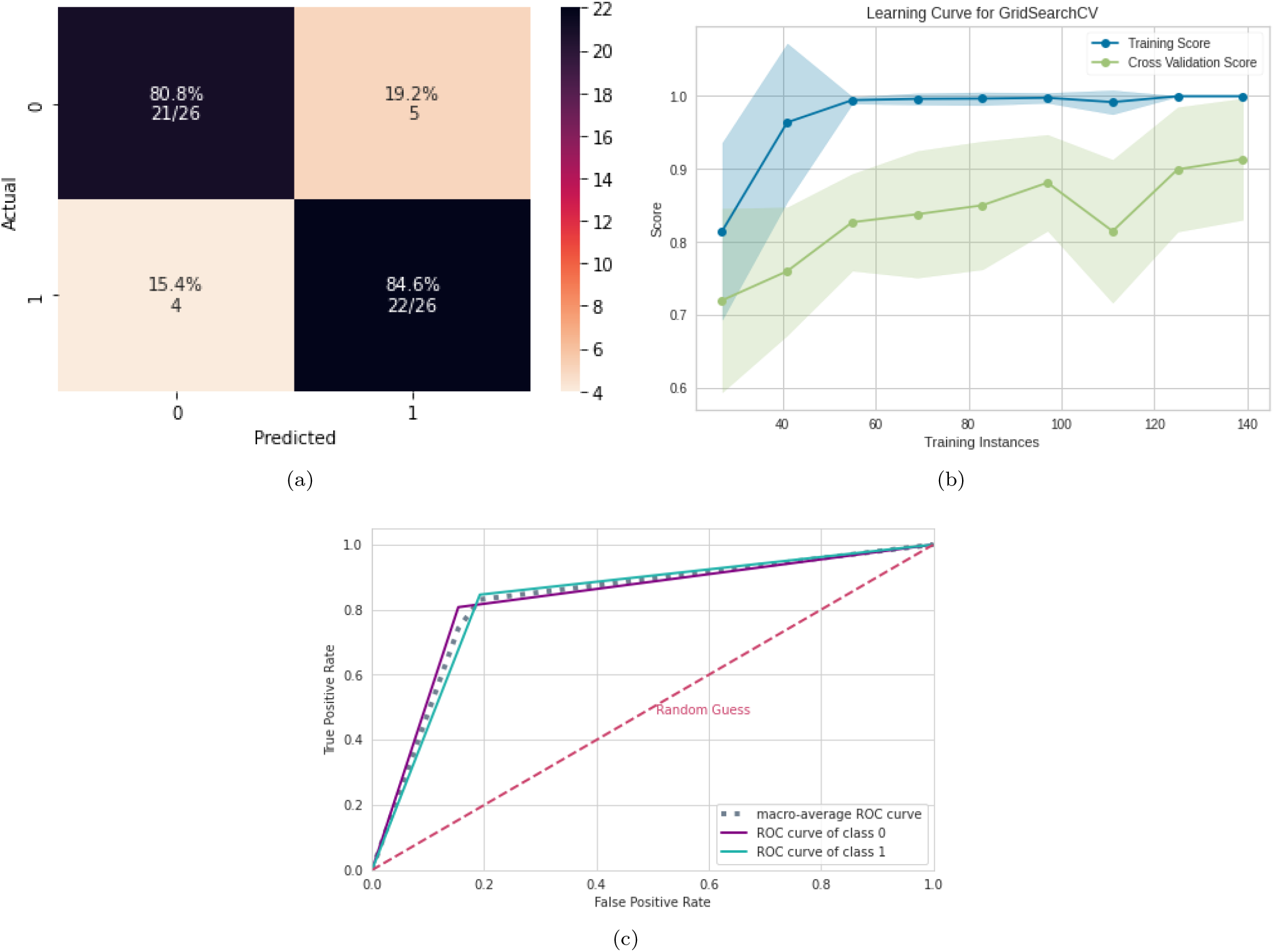
ML results using connectivity matrices. (a) Confusion matrix indicating a true negative rate of 80.8% (purple according to the color bar) and a true positive rate of 84.6% (blue according to the color bar). (b) Learning curve for the training accuracy (blue) and for test accuracy (green). (c) ROC curve with class 0 (control) and class 1 (after inhalation of DMT).

In order to reveal the importance of the connections between electrode pairs (brain connections) by considering the combination of the best EEG frequency bands (high alpha, low beta and delta), the SHAP values were calculated. The results are shown in Figure 4. Clearly, the most important connection was between electrodes TP8 (temporal and parietal region) and C3 (central region). In addition, the presentation of the data in Figure 4 shows that for the connection between TP8 and C3, low values of correlation (blue dots) were important for detecting the presence of DMT (positive SHAP values), and high values of correlation (red dots) were important for detecting the absence of DMT (negative SHAP values). The second most important connection was between the electrodes FC5 and P8. The corresponding brain regions are depicted in Figure 5.

**FIG. 4:**
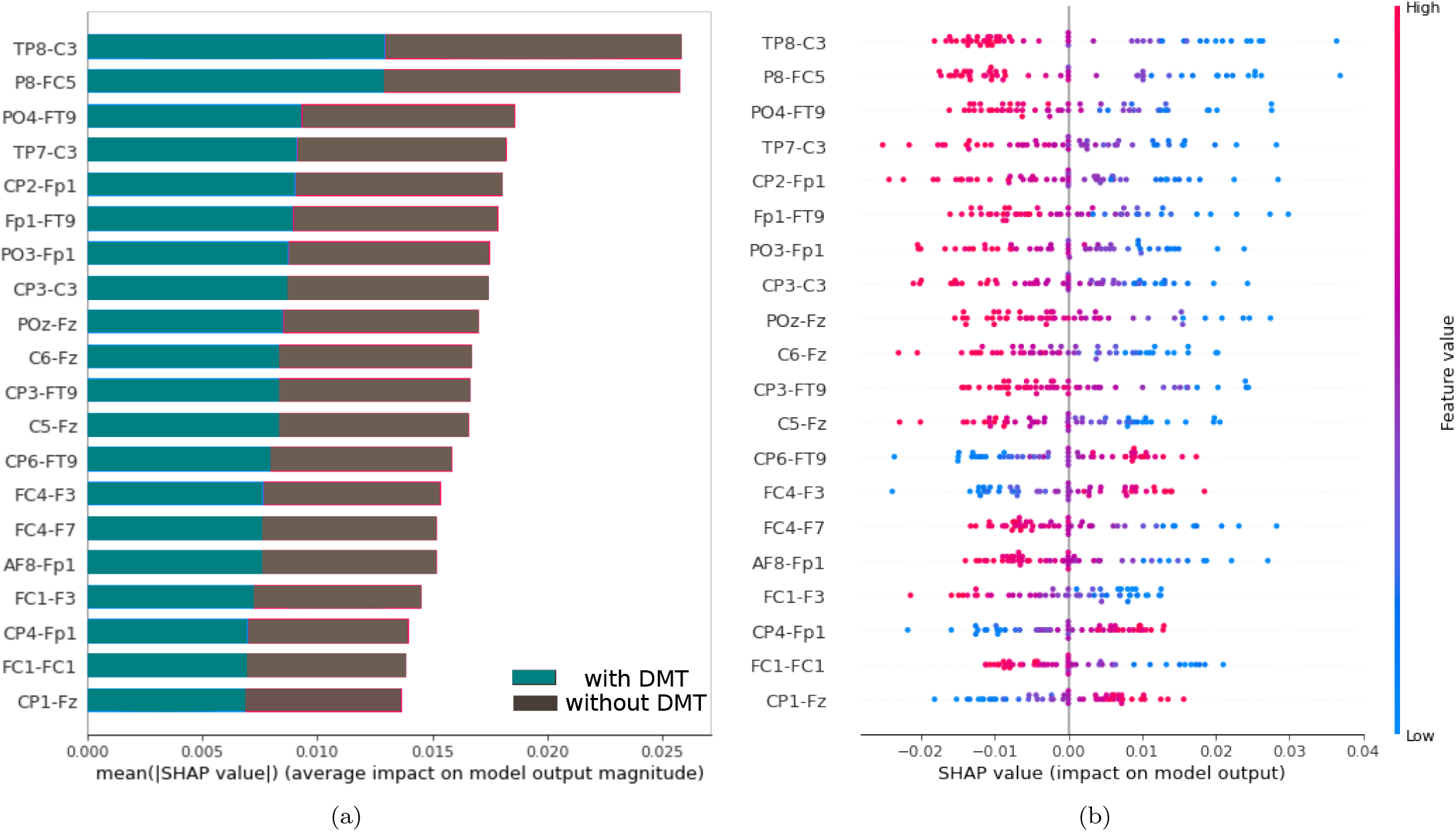
Feature importance ranking for the SVM classifier with electrode correlation (brain regions) ranked in descending order of importance by considering the combination of the best EEG frequency bands (high alpha, low beta and delta). The connection between the regions TP8 and C3 is the most important to classify the effect of DMT. (a) Feature ranking based on the average of absolute SHAP values over all subjects considering both classes (gray: control, cyan: after inhalation of DMT). (b) Same as (a), but additionally showing details of the impact of each feature on the model.

**FIG. 5:**
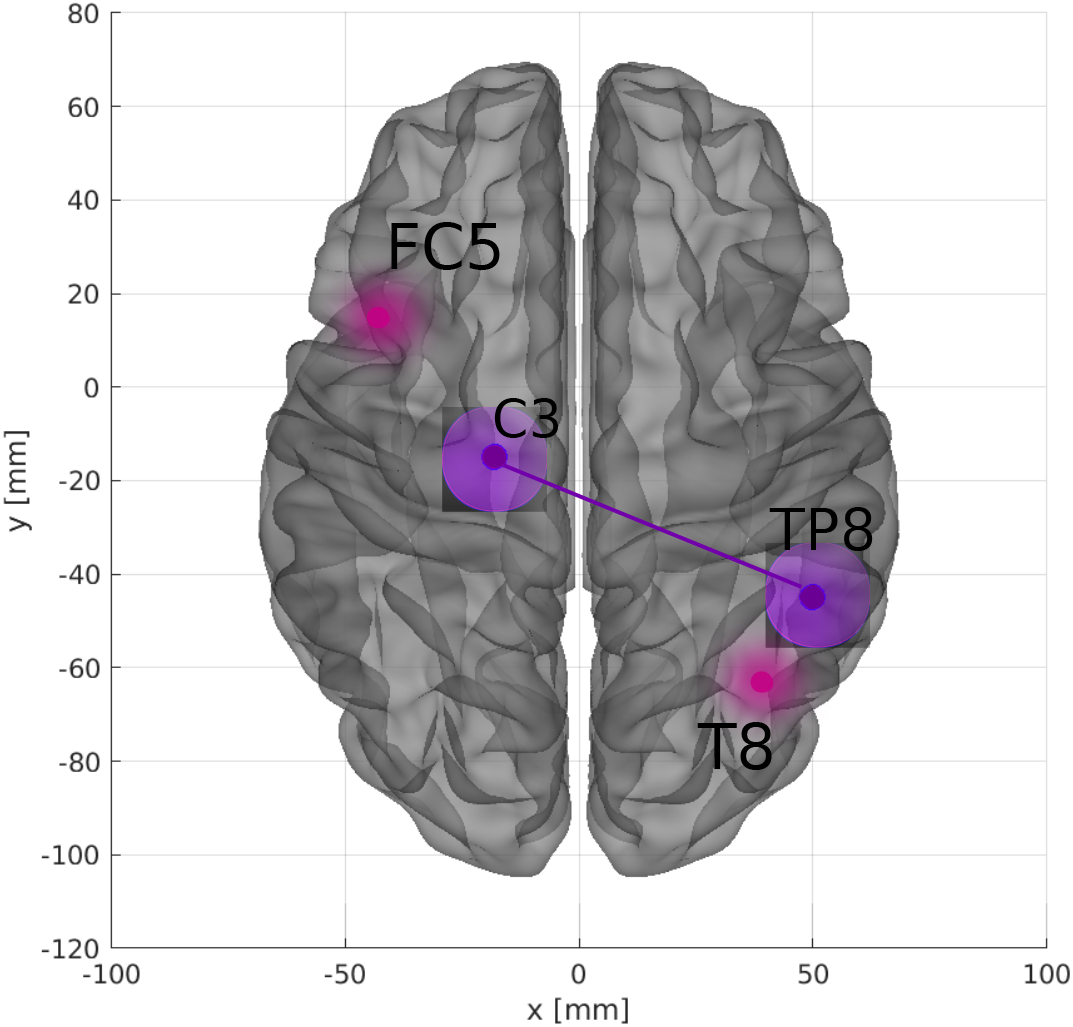
Brain plot displaying these electrode pairs which contributed most to the classification result based on connectivity matrices. The brain plot was made using Braph tool [150], based on the coordinates in [151, 152].

### B. Complex network measures

We received the best performance considering complex network measures for the delta frequency band (test sample performance with mean AUC of 0.89, mean precision of 0.91, mean F1 score of 0.88, mean recall of 0.88 and mean Acc. of 0.89), see Table I. The precision measure is related to the positive class (with DMT). Thus, since the precision was higher than the recall, we conclude that the model slightly better detects the presence of DMT than its absence. Furthermore, see appendix D for similarity of results obtained for each frequency.

In Figure 6, the confusion matrix (Figure 6 (a)), the learning curve (Figure 6 (b)), and the ROC curve (Figure 6 (c)) are plotted. Again, the entire database is necessary in order to get the highest accuracy. All the other results can be found in the Appendix C

**FIG. 6:**
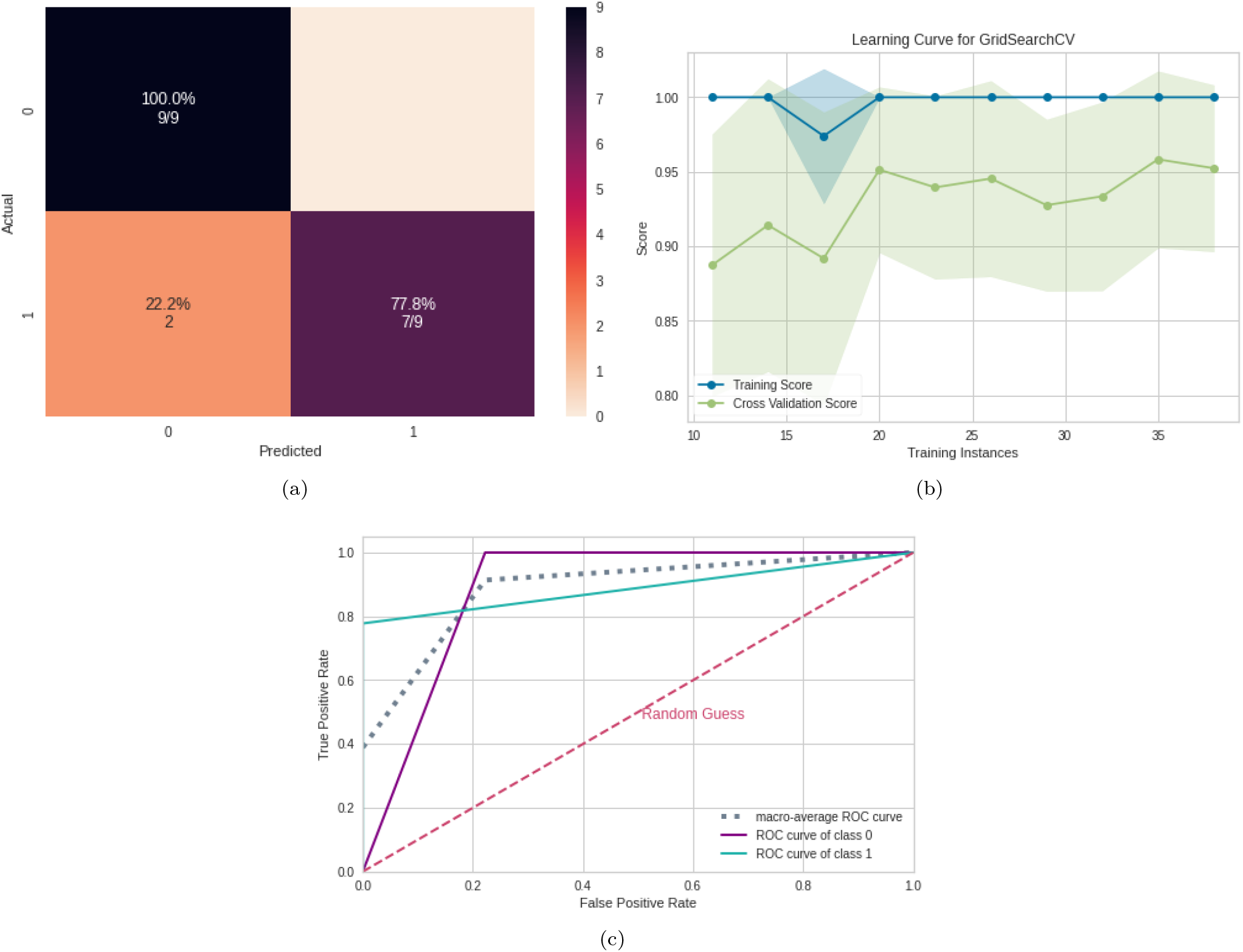
ML results using complex network measures. (a) Confusion matrix indicating a true negative rate of 77.8% (purple according to the color bar) and a true positive rate of 100.0% (blue according to the color bar). (b) Learning curve for the training accuracy (blue) and for test accuracy (green). (c) ROC curve with class 0 (control) and class 1 (after inhalation of DMT).

Based on the SHAP values in Figure 7 it can be seen that the most important measure for the model was the CCy, followed by the ALPC measure and the APL. In addition, high values of the CC measure (pink dots) indicate its importance for the detection of the absence of DMT (negative SHAP values), see Figure 7 (b).

**FIG. 7:**
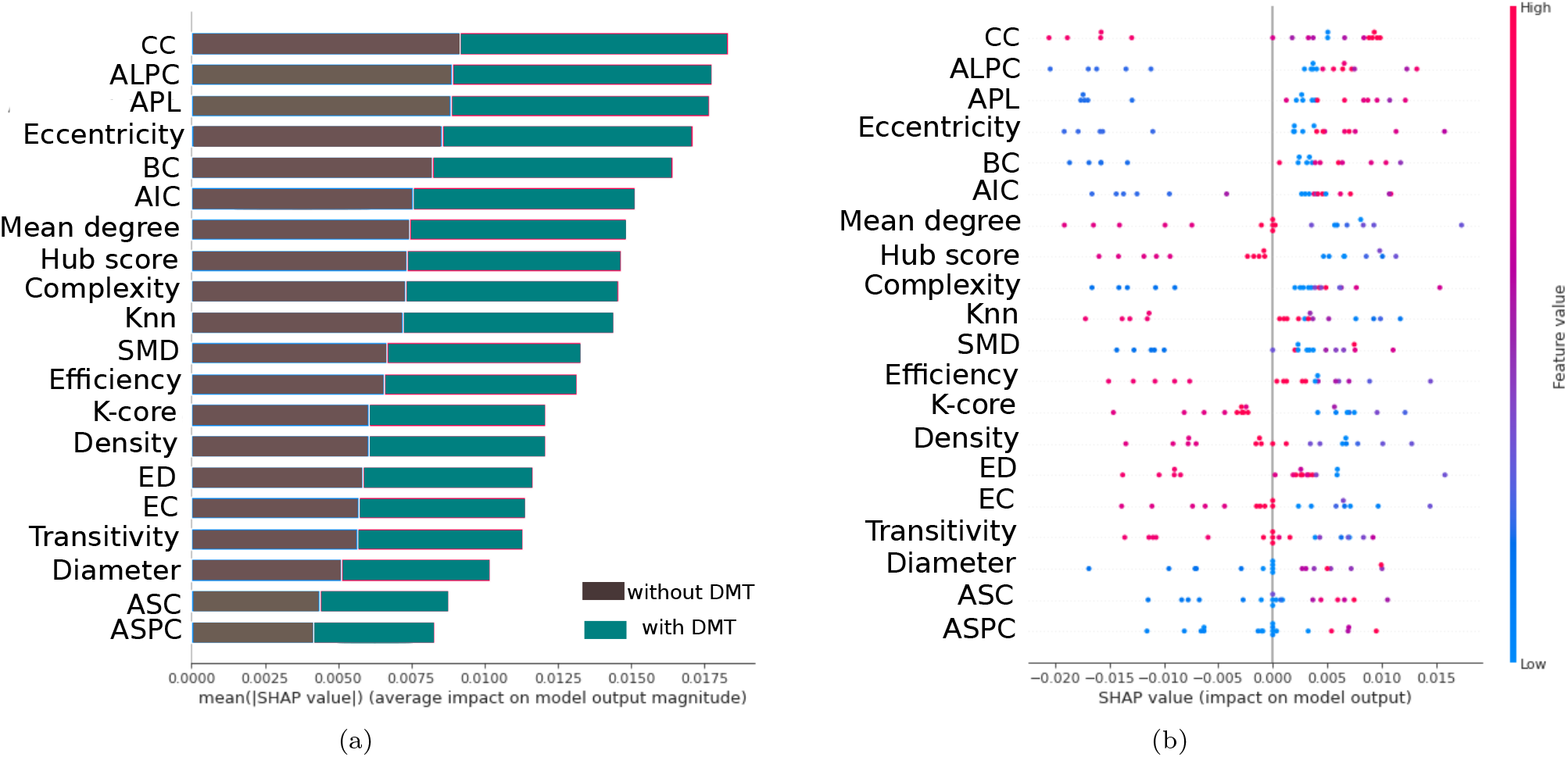
Feature importance ranking for SVM classifier with features ranked in descending order of importance. The CC measure is the most important to classify the effect of DMT. (a) Feature ranking based on the average of absolute SHAP values over all subjects considering both classes (gray: control, cyan: after inhalation of DMT). (b) Same as (a) but additionally showing details of the impact of each feature on the model.

## V. DISCUSSION

In the previous sections, we presented a computational workflow including data preprocessing and ML algorithm revealing acute differences in the brain activity before and after the consumption of the psychedelic drug DMT. As a result we achieved a classification accuracy of at least 82%. We further showed that the classification accuracy based on complex network measures (89%) was higher than that based on the connectivity matrix alone (see Table I), suggesting that these measures are important to capture differences in brain activity.

Further, we searched for descriptive parameters related to changes in the functional network structure by ranking the importance of features which contributed to the classification result. The results are discussed in this section with the aim to get insights into the effects of DMT consumption on the brain in terms of EEG frequency band (subsection V A), connection of the most activated brain regions (subsection V B), and measures of complex networks (subsection V C).

### A. Frequency bands

With our workflow, we were able to identify these frequency bands which were mostly modified after the intake of DMT. We found that classification results received with the connectivity matrices and the complex network measures are strongly based on acute changes in the delta band. Thus, changes in the delta band were most robust for both input data types.

This observation corresponds to what was also found in the literature [28, 74]. Delta band activity is usually associated with states where there is no wakefulness, such as sleep [153, 154] and coma [155]. However, some studies such in [156] observed that the delta frequency is present even when there are behavioral responses, such as in propofol anesthesia, postoperative delirium, and in powerful psychedelic states. Moreover, delta band activity has also been detected in studies involving spiritual experiences [157–159] and meditation states [160, 161].

Although the increase of delta band activity points clearly to an altered state of consciousness after the inhalation of DMT, also other frequency bands were affected. We found for the connectivity matrices that in addition to delta, high alpha and low beta bands were important features. This finding is supported by [74] who describes that inhalation of DMT reduces the alpha band activity while increasing the delta and gamma band at the same time [162]. According to the authors, the increase in gamma is associated with subjective perceptions typical of mystical experiences. In our data, we observed no changes in the gamma band.

### B. Connection between brain regions

With the connectivity matrices, we found that classification results are strongly based on a decreasing correlation between the temporal/parietal (TP8) and the central brain (C3) region after DMT uptake. These brain areas correspond to occipitotemporal (Right BA37), primary somatosensory cortex and the motor cortex (Left BA01/02), via Brodmann’s map [163]. The temporal lobe is associated with perception and production of speech, hearing, memory and emotional processes, because it is connected to the amygdala and the limbic system [145]. The right temporal region, TP8 found here, is associated with the recognition of familiar faces, with participation from the frontal cortex [164]. Furthermore, in humans, TP8 region contributes to the global processing of visual information [165]. The connection between this two regions, TP8 and C3, is involved in visual and tactile perceptions, and finger movements [166, 167].

In addition, the correlation between the electrodes FC5 (frontal region) and P8 (parietal region) contributed significantly to the classification result. These regions correspond to Left BA6 and Right BA19 of Brodmann’s map, respectively. The frontal region is involved in cognitive processing, planning behavior, and has connections to the somatosensory cortex, motor, and auditory areas [168, 169] and limbic system, and is also involved in emotions. The placement of FC5 electrode encompasses the region of the precentral gyrus in the pre motor region, which is responsible for controlling voluntary motor movement of the body. This region also includes a portion of the supplementary motor cortex, responsible for planning the voluntary movement of the limbs [170]. The P8 region, on the other hand, is located in the lateral occipital cortex, responsible for integrating different types of information so that our interaction with the environment is efficient, forming representational spaces through perception, semantics, through perception, semantics, and motor functions [171].

Studies using other psychedelics, such as LSD, have found reduced functional connectivity in the anterior medial prefrontal cortex, and time-specific effects were correlated with different aspects of subjective experiences under the effect of psychedelics [172]. Psilocybin consumption, on the other hand, was related to decreased functional connectivity between the medial temporal lobe and high-level cortical regions. The changes found in the cortical regions reported above, are related to the visual, sensory, perceptual, and motor type experiences experienced by the volunteers during the during the use of these two psychedelics (LSD and Psilocybin). Correlating our findings with previous studies, the FC5 and P8 regions also found here are part of the cortical region and a possible inference is that they are related to the participants’ experience with DMT in [74] in which it was reported that 13 of 35 participants (equivalent to 37%) accessed a complete mystical experience ^3^. Although these two cortical regions, FC5 and P8, may be related to the participants’ sensory and visual experiences when using DMT, no interpretation of the connectivity between these two regions has been obtained, since there is no information in the literature.

### C. Complex network measures

Concerning the measures of complex networks, the most important was the CC. CC is a centrality measure which is defined as the inverse of the average length of the shortest path from one node to all other nodes in the network [173]. The idea is that important nodes participate in many shortest paths within a network and, therefore, play an important role in the flow of information in the brain [94]. ALPC was the second important measure, which is associated to the size of the largest community found by the label propagation community (LPC) detection algorithm. Increased values (compared to controls) of this metric are associated with the effect of DMT (see Figure 7 (b)) indicating communities with increased average path lengths after the use of DMT, in other words, larger communities. The third important metric was the APL which is the average of all shortest paths. The shortest path *d*_*ij*_ (also known as the geodesic path) between two nodes *i* and *j*, is defined as the shortest of all possible paths between these vertices. Increased values for APL were associated with the presence of DMT (Figure 7(b)).

In the brain of large vertebrates there are two contrasting concepts: functional segregation (or specialization) and integration (or distributed processes) [174]. Anatomical and functional segregation refers to the existence of specialized neurons and brain areas organized in modules [175] which correspond to communities where their members have high connectivity among themselves and few connections with members of other modules [176]. As opposed to segregation, neuron units don’t operate in isolation [175], there are regions of the brain (distributed system of the cerebral cortex) capable of combining specialized information, characterizing the concept of integration [173]. These regions have an executing function, benefiting from a high global efficiency of information transfer throughout the entire network [177]. The fact that we found larger communities and a longer average path with the use of DMT and the opposite in its absence, indicates a decrease of the brain integration, which might slow down the distribution of information. Larger brain communities were also found in [44] after the use of ayahuasca, a mixture containing DMT.

Furthermore, when looking at the transitivity, which is a measure of the propensity of nodes to be grouped together, and efficiency measure, which is a measure of how effective the exchange of information within a network is, both also presented in the rank of the most important measures for the model in Figure 7 (b), the presence of DMT decrease the values of these two measures. The transitivity is a measure of the efficiency of information transfer between all pairs of nodes in the graph [178] and a higher value of these measures indicates greater segregation [179]. On other hand, higher values of efficiency indicates greater integration of networks. Thus, we can infer that the integration and segregation decreased with the use of the DMT considering the delta frequency. A decrease in brain segregation has been found in studies using other psychedelics such as LSD [42, 172, 180]. Specifically in [180], the authors concluded that the use of LSD caused a decrease in the integration and segregation of brain networks, supporting the idea that cortical brain activity becomes more “entropic” under psychedelics [181]. However, as pointed out in [42], psychedelics not only render the brain more random, but with normal organization disruption, they also produce strong functional and topologically far-reaching connections not seen in the normal state. Thus, even though our results show that integration and segregation have been disrupted, further experiments should be conducted to verify if there have been new long-distance connections as shown in the literature.

## VI. CONCLUSION AND FUTURE WORK

In summary, our results demonstrated that the application of ML methods was able to automatically reveal changes in brain functional connectivity induced by DMT consumption considering a two-class classification based on (A) the connectivity matrix or (B) complex network measures. The workflow developed here was indeed powerful for detecting the brain changes caused due to the psychedelic substance, with case (B) showing the highest AUC (89%), indicating that complex network measurements best capture the brain changes that occur due to DMT use. In terms of frequency, the workflow employed here detected that the delta frequency was most associated with DMT. Although DMT induces an altered state of consciousness with the presence of delta, other frequencies were important for recognizing the pattern of brain activity with the use of this substance, such as high alpha and low beta, through the connectivity matrix. This may suggest that the combination between the brain frequencies may represent an important point to be investigated, to further define the altered state of consciousness induced by DMT.

Furthermore, by using the SHAP value it was possible to interpret the results of the ML algorithms with a biological interpretation associated with the use of DMT on EEG data. The most important connections found with the use of DMT were between the temporal (TP8) and central cortex (C3) regions, followed by the connection between the precentral gyrus (FC5) and the lateral occipital cortex (T8). The connection between regions TP8 and C3 has been found in the literature associated with finger movements that might have occurred during DMT consumption. However, the connection between cortical regions FC5 and P8 has not been found in the literature and is presumably related to emotional, visual, sensory, perceptual, and mystical experiences of the volunteers during DMT consumption.

Concerning the measures of complex networks similar to what was found with the use of ayahuasca in [75], the most important was the centrality measure CC. Also, the fact that we found larger communities and a longer average path with the use of DMT and the opposite in its absence, indicates that this balance between functional segregation and integration was disrupted. This suggests that the distribution of information is slower. This findings supports the idea that cortical brain activity becomes more entropic under psychedelics. However, from the literature, psychedelics don’t simply make the brain more random, but after the normal organization is disrupted, strong and topologically far-reaching functional connections emerge that are not present in the normal state. Therefore, we would like to investigate in the long term how psychedelics change the functional connectivity of the brain using our workflow. Overall, a robust computational workflow has been developed here with an interpretability of how DMT (or other psychedelics) modify brain networks and insights into their mechanism of action. Finally, the same methodology applied here may be useful in interpreting EEG time series from patients who consumed other psychedelic drugs and can help obtain a detailed understanding of functional changes in the neural network of the brain as a result of drug administration. Thus, in future work we intend to use this methodology on the psychedelic drug called ketamine [182].

## Data Availability

All data are available online at https://zenodo.org/record/3992359

## VII. ACKNOWLEDGEMENTS

F.A.R. acknowledges CNPq (grant 309266/2019-0) and FAPESP (grant 19/23293-0) for the financial support given for this research. T.G.L.O.T acknowledges FAPESB (grant number 307/2020 – Cota 2020; BOL0202/2020) for the financial support given this research. A.M.P. acknowledges FAPESP (grant 2019/22277-0) for the financial support given this research.

## Appendix A

### Grid search hyperparameter tuning

The Figure 8 contains the values used in the present work where for one of the models (considering all frequencies and comparation of dmt and open eyes control) the combination of hyperparameter values and the grid search was plotted in relation to the AUC metric.

**FIG. 8:**
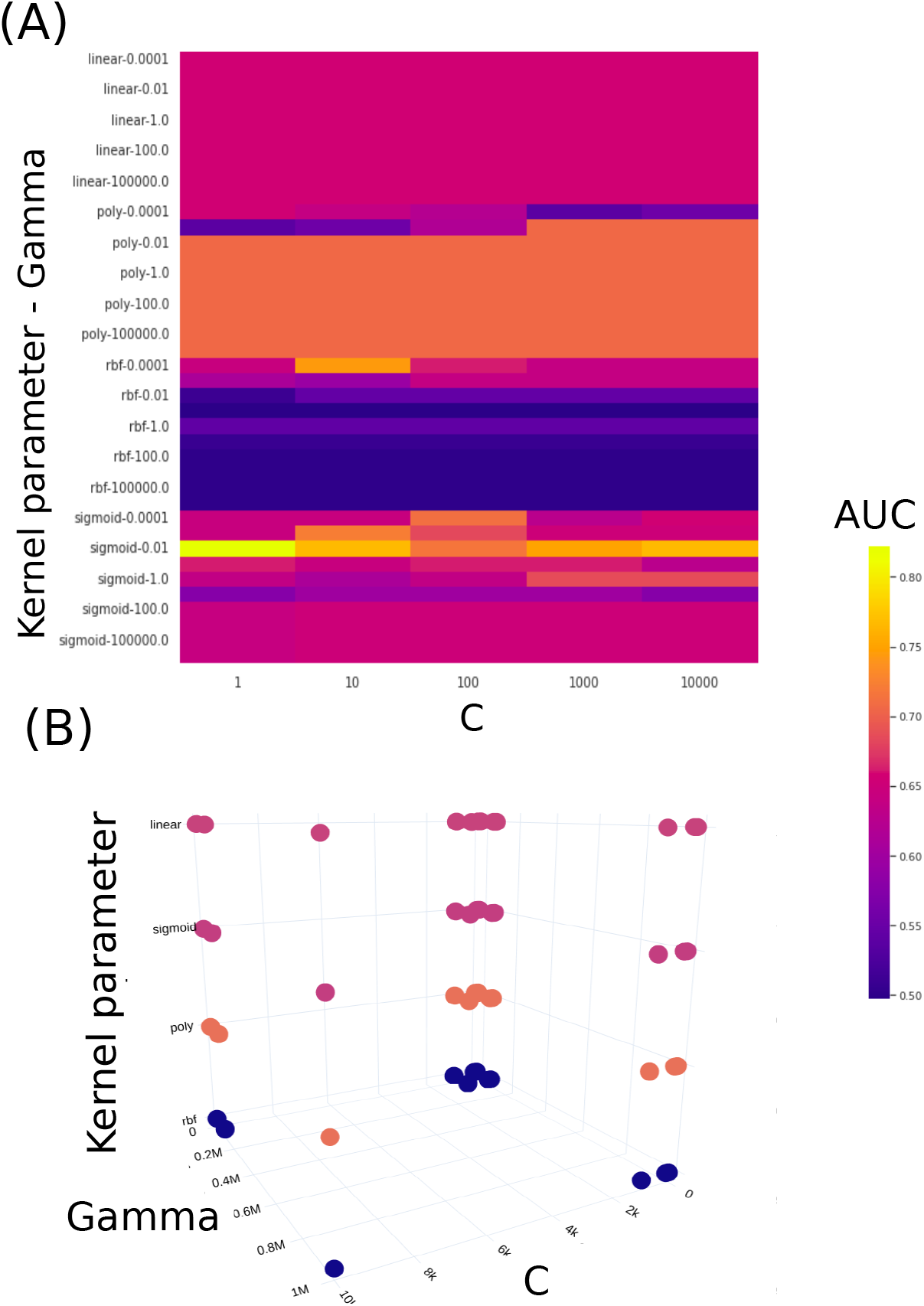
Figure containing the values of each hyperparameter of the SVM varied with the grid search. In (A) for the model considering all frequencies and comparing the subject DMT and without DMT with the eye closed, the two-dimensional plot with the x-axis being the values of the parameter C and the y-axis being the values of the kernel and gamma function. For each combination of values and hyperparameters, AUC performance was obtained (whose obtained values are illustrated in the color table). In (B), the three-dimensional plot of (A) in which each hyperparameter corresponds to an axis.

## Appendix B

### Results comparing different band frequencies

Appendix B: Results comparing different band frequencies
| Band frequencies | Subset | AUC | Acc. | F1 score | Recall | Precision |
| --- | --- | --- | --- | --- | --- | --- |
| All frequencies | Train | 0.90 | 0.90 | 0.90 | 0.90 | 0.90 |
|  | Test | 0.67 | 0.67 | 0.67 | 0.67 | 0.68 |
| Low alpha | Train | 1.00 | 1.00 | 1.00 | 1.00 | 1.00 |
|  | Test | 0.67 | 0.67 | 0.67 | 0.67 | 0.67 |
| High alpha | Train | **1.00** | **1.00** | **1.00** | **1.00** | **1.00** |
|  | Test | **0.72** | **0.72** | **0.72** | **0.72** | **0.73** |
| Low beta | Train | **0.98** | **0.98** | **0.98** | **0.98** | **0.98** |
|  | Test | **0.78** | **0.78** | **0.78** | **0.78** | **0.78** |
| Mid beta | Train | 0.98 | 0.98 | 0.97 | 0.98 | 0.98 |
|  | Test | 0.45 | 0.47 | 0.40 | 0.45 | 0.42 |
| High beta | Train | 1.00 | 1.00 | 1.00 | 1.00 | 1.00 |
|  | Test | 0.56 | 0.56 | 0.45 | 0.56 | 0.76 |
| Gamma | Train | 0.67 | 0.67 | 0.67 | 0.67 | 0.67 |
|  | Test | 0.44 | 0.44 | 0.42 | 0.44 | 0.43 |
| Delta | Train | **1.00** | **1.00** | **1.00** | **1.00** | **1.00** |
|  | Test | **0.72** | **0.72** | **0.72** | **0.72** | **0.73** |
| Theta | Train | 0.61 | 0.61 | 0.61 | 0.61 | 0.61 |
|  | Test | 0.50 | 0.50 | 0.50 | 0.50 | 0.50 |

## Appendix C

### Results considering complex network measures and different frequencies band

Appendix C: Results considering complex network measures and different frequencies band
| Band frequencies |  | AUC | Acc. | F1 score | Recall | Precision |
| --- | --- | --- | --- | --- | --- | --- |
| All frequencies | Train | 1.00 | 1.00 | 1.00 | 1.00 | 1.00 |
|  | Test | 0.69 | 0.69 | 0.68 | 0.69 | 0.73 |
| Low alpha | Train | 1.00 | 1.00 | 1.00 | 1.00 | 1.00 |
|  | Test | 0.72 | 0.72 | 0.70 | 0.72 | 0.82 |
| High alpha | Train | 1.00 | 1.00 | 1.00 | 1.00 | 1.00 |
|  | Test | 0.61 | 0.61 | 0.54 | 0.61 | 0.76 |
| Low beta | Train | 1.00 | 1.00 | 1.00 | 1.00 | 1.00 |
|  | Test | 0.56 | 0.56 | 0.56 | 0.56 | 0.56 |
| Mid beta | Train | 0.98 | 0.98 | 0.98 | 0.98 | 0.98 |
|  | Test | 0.45 | 0.47 | 0.40 | 0.45 | 0.42 |
| High beta | Train | 1.00 | 1.00 | 1.00 | 1.00 | 1.00 |
|  | Test | 0.56 | 0.56 | 0.45 | 0.56 | 0.76 |
| Gamma | Train | 1.00 | 1.00 | 1.00 | 1.00 | 1.00 |
|  | Test | 0.17 | 0.17 | 0.14 | 0.17 | 0.13 |
| Delta | Train | **1.00** | **1.00** | **1.00** | **1.00** | **1.00** |
|  | Test | **0.89** | **0.89** | **0.88** | **0.88** | **0.91** |
| Theta | Train | 1.00 | 1.00 | 1.00 | 1.00 | 1.00 |
|  | Test | 0.50 | 0.50 | 0.33 | 0.50 | 0.25 |

## Appendix D

### Similarity of results obtained for each frequency

The SHAP value calculated for each frequency band was also considered. For each band a vector of connection between electrode pairs and its respective SHAP value found by the model is generated. For each of these vectors the Euclidean distance between them is then calculated generating a distance matrix of these vectors. This aims to quantify how close resulting vectors are. The distance matrix is displayed in form of a cluster map, see Figure 9, where vectors with a distance less than 0.2 are connected hierarchically in a dendrogram indicating clusters. Here, the cluster is most prominent between the low beta and the delta frequency band vector.

**FIG. 9:**
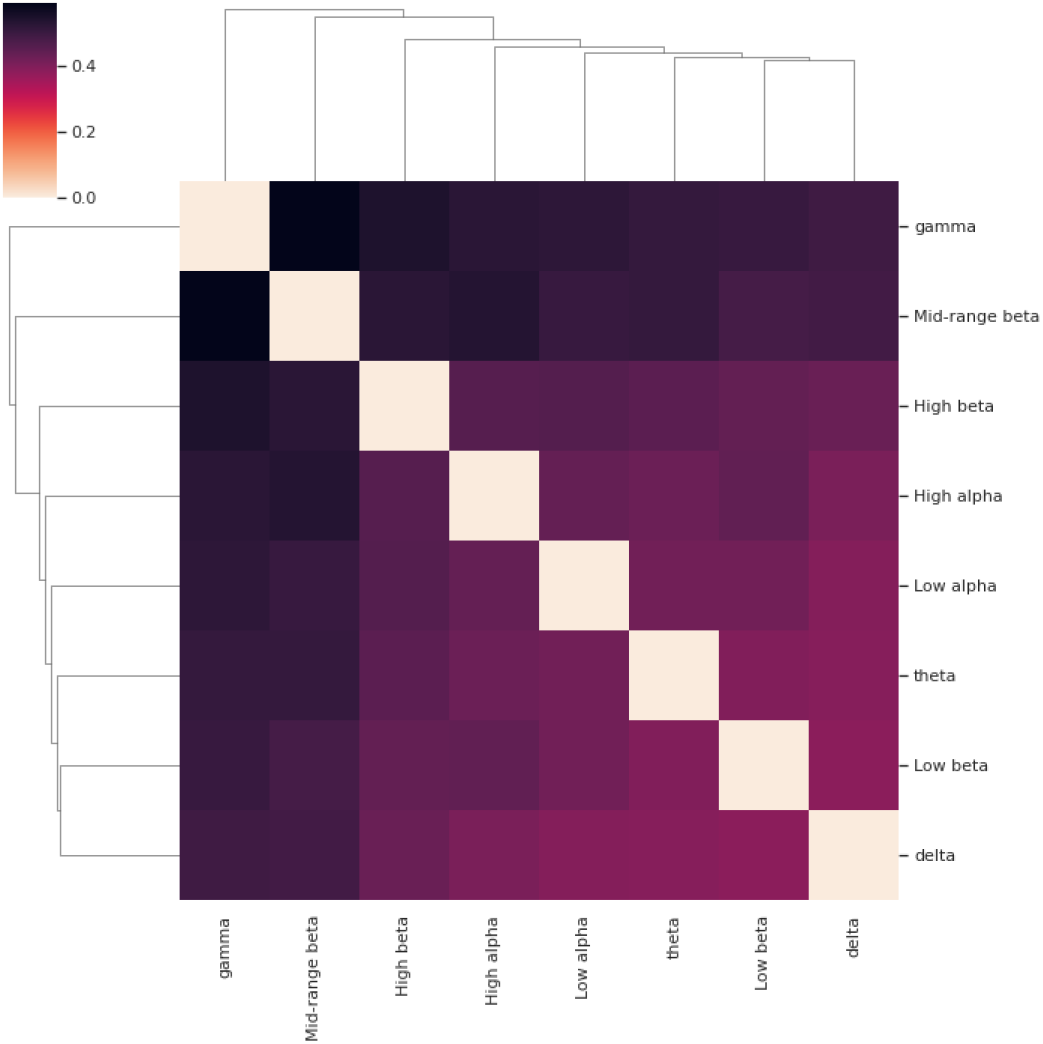
Cluster map with the Euclidean distance of each band SHAP value vector. The delta bands and low beta frequency are the closest frequencies, forming in the cluster map a connection.

The same was made for the complex network measure and, then, a cluster map with the Euclidean distance between vectors containing SHAP values for each complex network measure is generated (see Figure 10). All vectors, except low beta, are very close to each other. This proximity indicates that the results obtained were similar, with other words the connections between the electrodes and their respective SHAP value were similar for all frequency bands. Thus, it can be seen that the results of the SHAP value vectors of each frequency, with the exception of the low beta frequency, were very close, which means that they show a great similarity between them.

**FIG. 10:**
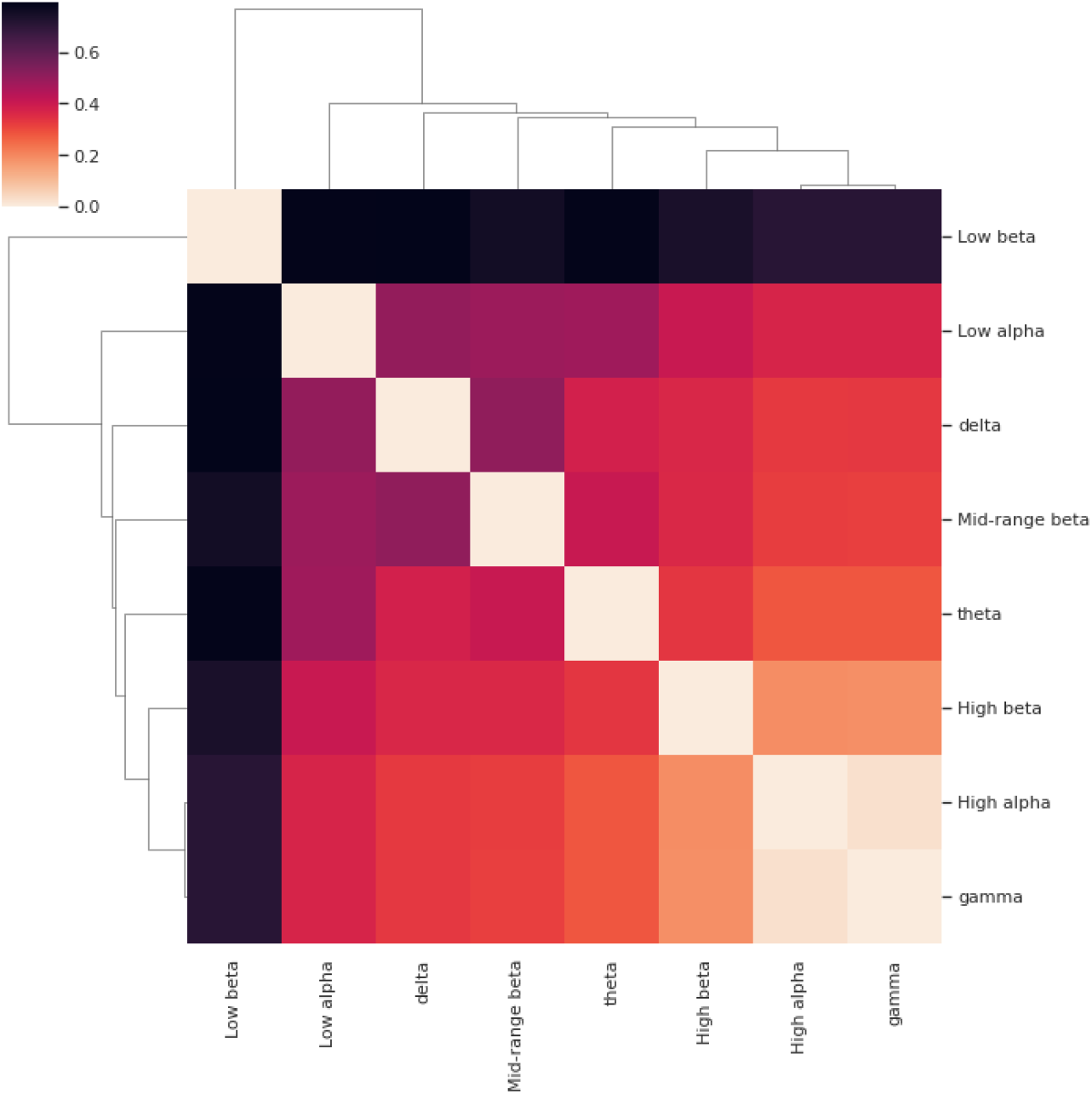
Cluster map showing the Euclidean distance of the SHAP value vectors for different frequency bands. Frequency band vectors, except low beta, are very close to each other, indicating great similarity.

## Footnotes

1 The network nodes can be representations from of neurons (¡1*µm*, microscale) to brain regions (*≈* 10 cm, macro scale)

2 Avaiable on Zenodo. https://doi.org/10.5281/zenodo.3992359

3 According to [74], there was a significant positive correlation between items of the following scales, used with the volunteers in this study: the 5D altered states of consciousness scale (5D-ASC), mystical experience scales such as the affective component of the NDE scale and MEQ-30, and the social interactions on the psychedelic experience (post-experience questionnaire). Among the experiences accessed by the volunteers during the use of DMT, the scores with the highest percentages (mean; SD) for the 5D-ASC scale were: elementary imagery (85.27%; 20.72%), blissful (61.77%; 25.9%), complex imagery (50.21%;20.4%), spiritual (49.61%; SD 29%) and disembodiment (47.58%; 31.3%). For the NDE scale, related to positive mood, it was 60.94% (26.39%) for affect experience. For MEQ-30 scale, was found awe, with 46.29% (14.57) and for post-experience questionnaire scale, setting (76.48%; 25.38) and social (61.79; 3.91%)

